# The Estimations of the COVID-19 Incubation Period: A Scoping Reviews of the Literature

**DOI:** 10.1101/2020.05.20.20108340

**Authors:** Nazar Zaki, Elfadil A. Mohamed

## Abstract

**Background:** A novel coronavirus (COVID-19) has taken the world by storm. The disease has spread very swiftly worldwide. A timely clue which includes the estimation of the incubation period among COVID-19 patients can allow governments and healthcare authorities to act accordingly.

**Objectives:** to undertake a review and critical appraisal of all published/preprint reports that offer an estimation of incubation periods for COVID-19.

**Eligibility criteria:** This research looked for all relevant published articles between the dates of December 1, 2019, and April 25, 2020, i.e. those that were related to the COVID-19 incubation period. Papers were included if they were written in English, and involved human participants. Papers were excluded if they were not original (e.g. reviews, editorials, letters, commentaries, or duplications).

**Sources of evidence:** COVID-19 Open Research Dataset supplied by Georgetown’s Centre for Security and Emerging Technology as well as PubMed and Embase via Arxiv, medRxiv, and bioRxiv.

**Charting methods:** A data-charting form was jointly developed by the two reviewers (NZ and EA), to determine which variables to extract. The two reviewers independently charted the data, discussed the results, and updated the data-charting form.

**Results and conclusions:** screening was undertaken 44,000 articles with a final selection of 25 studies referring to 18 different experimental projects related to the estimation of the incubation period of COVID-19. The majority of extant published estimates offer empirical evidence showing that the incubation period for the virus is a mean of 7.8 days, with a median of 5.01 days, which falls into the ranges proposed by the WHO (0 to 14 days) and the ECDC (2 to 12 days). Nevertheless, a number of authors proposed that quarantine time should be a minimum of 14 days and that for estimates of mortality risks a median time delay of 13 days between illness and mortality should be under consideration. It is unclear as to whether any correlation exists between the age of patients and the length of time they incubate the virus.

## 1. Introduction

At the start of 2020, a new form of coronavirus (COVID-19) was found to be the source of infection responsible for an epidemic of viral pneumonia in Wuhan, China, a region in which the first patients began to show symptoms in December 2019. At the time of writing (July 28, 2020) over 16.5 million people globally have caught the virus, of whom more than 650,000 have died. The novel virus, causing severe acute respiratory disease, is thought to be from the same family as Middle East Respiratory Syndrome (MERS) coronavirus and Severe Acute Respiratory Syndrome (SARS) coronavirus, but it is unique in its own right. This means that central epidemiological parameters, which include the incubation period, are being urgently researched in real-time from case reports while the epidemic is continuing [1]. The incubation period of a virus represents the time span from the probable earliest contact with a source of transmission and the earliest recognition of the first symptoms. Accurately estimating the length of the incubation period is essential for effective contemporary public health measures to be taken [2]. If health authorities know what the incubation period is then they will know for how long a healthy individual has to be monitored and have their movement restricted (quarantine period) [3]. Correctly estimating the incubation period will also help us to comprehend how infectious COVID-19 is, make estimations of the size of the pandemic, and decide on the best course of action [4]-[7]. With insufficient data available to definitively state what the incubation period for this virus is, the World Health Organisation (WHO) is working with a broad range of 0-14 days, the European Centre for Disease Prevention and Control (CDCDC) is working with a range of 2-14 days, and a number of studies have made the assumption that the incubation period is similar to that of the MERS and SARS coronaviruses [1].

Various infectious/viral diseases have a variety of incubation periods. Nevertheless, for some infectious diseases, we are relatively certain about the incubation period. For every individual, the level of pathogens invading the body, their ability to reproduce and resist treatment will differ, and so for any specific disease, the data related to incubation periods should be treated with logarithm normal distribution [8]. Log-normal distribution represents the continuous probability distribution for a random variable with normal logarithm distribution. Incubation periods can usually be measured via biological experimentation and physiological observation. Accurately determining the incubation period will significantly influence controls to prevent transmission of the disease and official policy regarding it. Nevertheless, determining the incubation period for COVID-19 is no simple matter, due to the fact that there is no consistency in the quality of the available data. One reason for this is that generally, we can only discover the times when the patient was in contact with persons carrying the virus, and then assume that the incubation period runs from the earliest date of exposure to the appearance of clinical symptoms or medical diagnosis. This way of calculating the incubation period may well be responsible for overestimation. The objective of this study is to represent a Scoping Reviews of the literature in order to answer the essential question of what length the COVID-19 incubation period is. Due to the fact that no method currently exists, that can make an accurate estimation of the incubation period, the only option is to draw lessons from past experience/practice.

## 2. Methodology

### 2.1. Protocol and registration

The protocol in this paper was drafted using the Preferred Reporting Items for Systematic Reviews and Meta-analysis extension for Scoping Reviews (PRISMA-ScR) [32]. The Scoping Reviews is a type of knowledge synthesis, follow a systematic approach to map pieces of evidence, identify the main concepts behind a topic; and determine where the gaps are.

### 2.2. Eligibility criteria

This research looked for all relevant published articles between the dates of December 1, 2019, and April 25, 2020, i.e. those that were related to the COVID-19 incubation period. Papers were included if they were written in English, and involved human participants. Papers published via Arxiv, medRxiv, and bioRxiv were also considered. Papers were excluded if they were not original (e.g. reviews, editorials, letters, commentaries, or duplications).

### 2.3. Information sources

A search was run with the “COVID-19 Open Research Base set” provided by Georgetown’s Centre for Security and Emerging Technology. This dataset can be found at https://cset.georgetown.edu/covid-19-open-research-dataset-cord-19/. It comprises more than 55,000 academic articles, and more than 44,000 of them have some reference to COVID-19, SARS-CoV-2, and other forms of coronavirus. This dataset is openly available to the global research community to employ using new techniques in Natural Language Processing (NLP) and other forms of Artificial Intelligence for the generation of novel insights supporting the continuing battle with the pandemic. This dataset is regularly updated when new research appears in peer-reviewed publications and archive services, e.g. [bioRxiv](https://www.biorxiv.org/), [medRxiv](https://www.medrxiv.org/), et cetera.

### 2.4. Search

For reviewing every article in the dataset we employed the Bidirectional Encoder Representations from Transformers for Biomedical Text Mining (BioBERT) model [9], which is a biomedical language representation model created to assist in mining biomedical texts. The inspiration for the model comes from the pre-trained language model BERT [10] created by Devlin J.et al. BioBERT resolves the difficulties caused by moving from a trained corpus for general use to biomedical use and assists in the understanding of complex biomedical texts as are found with the work on COVID-19. The model was taken from the Github https://github.com/dmis-lab/biobert. Filtering of the articles was then undertaken using keywords and questions, e.g. “What is the incubation period of COVID/normal coronavirus/SARS-CoV-2/nCoV”.

### 2.5. Selection of sources of evidence

Besides the automated search conducted using the (BioBERT) model [9], an independent search was undertaken by the two reviewers (NZ and EA), who examined the PubMed, and Google Scholar databases. To increase consistency, both reviewers screened the automated and manually extracted publications and discussed the results before beginning screening for the review. Both reviewers independently evaluated the titles, abstracts, and then the full text of all publications identified by our automated and manual searches for potentially relevant publications. Disagreements on study selection and data extraction were settled by consensus and discussion. No third opinion was needed.

### 2.6. Data charting process

A data-charting form was jointly developed by the two reviewers (NZ and EA), to determine which variables to extract. The two reviewers independently charted the data, discussed the results, and continuously updated the data-charting form. The charting table was further refined at the review stage and updated accordingly.

### 2.7. Data items

We abstracted data on paper reference number, author name, publication venue, the incubation period reported, methods for estimating the incubation period, number of cases (patients), the location where the study is conducted, and additional note.

### 2.8. Critical appraisal of individual sources of evidence

We performed an in-depth assessment of each published/preprint report that offers an estimation of incubation periods among COVID-19 patients. Unlike other topics, the estimation of incubation periods for novel COVID-19 disease is specific and doesn’t need the usage of tools to appraise the quality of the knowledge synthesis methods.

### 2.9. Synthesis of results

We grouped the selected studies by the types of studies in which the estimations of incubation periods are mentioned which includes: Estimating the incubation period, transmission characteristics, clinical characteristics, epidemiological characteristics, and case studies. Where we identified a systematic review, we counted the number of studies included in the review that potentially met our inclusion criteria and noted how many studies were excluded and the reasons for the exclusions.

## 3. Results

### 3.1. Selection of sources of evidence

As illustrated in Fig. 1, a systematic approach was used to automatically screen the full “COVID-19 Open Research Dataset”. Through employing BioBERT, out of the 44,000 articles, just 25 articles were found that scored highly on confidence rating. An additional search was run employing Google Scholar and PubMed to make sure that every relevant article was included. Every paper previously found was in the top 25 results alongside four other articles. Each of the 29 papers has been read thoroughly, causing nine articles to be excluded from the list for the following reasons: one was simply a report of earlier studies into incubation periods, one related to techniques for estimating incubation periods, two were letters to editors, one was a literature review regarding the geometric features of SARS-CoV-2, one was a perspective paper, and the other four were either irrelevant or redundant. The ultimate list of 18 articles was split into five categories, with four articles focusing on the study of incubation periods, seven on characteristics of transmission, two on clinical characteristics, four case studies, and one last paper related to epidemiological characteristics.

**Fig. 1:**
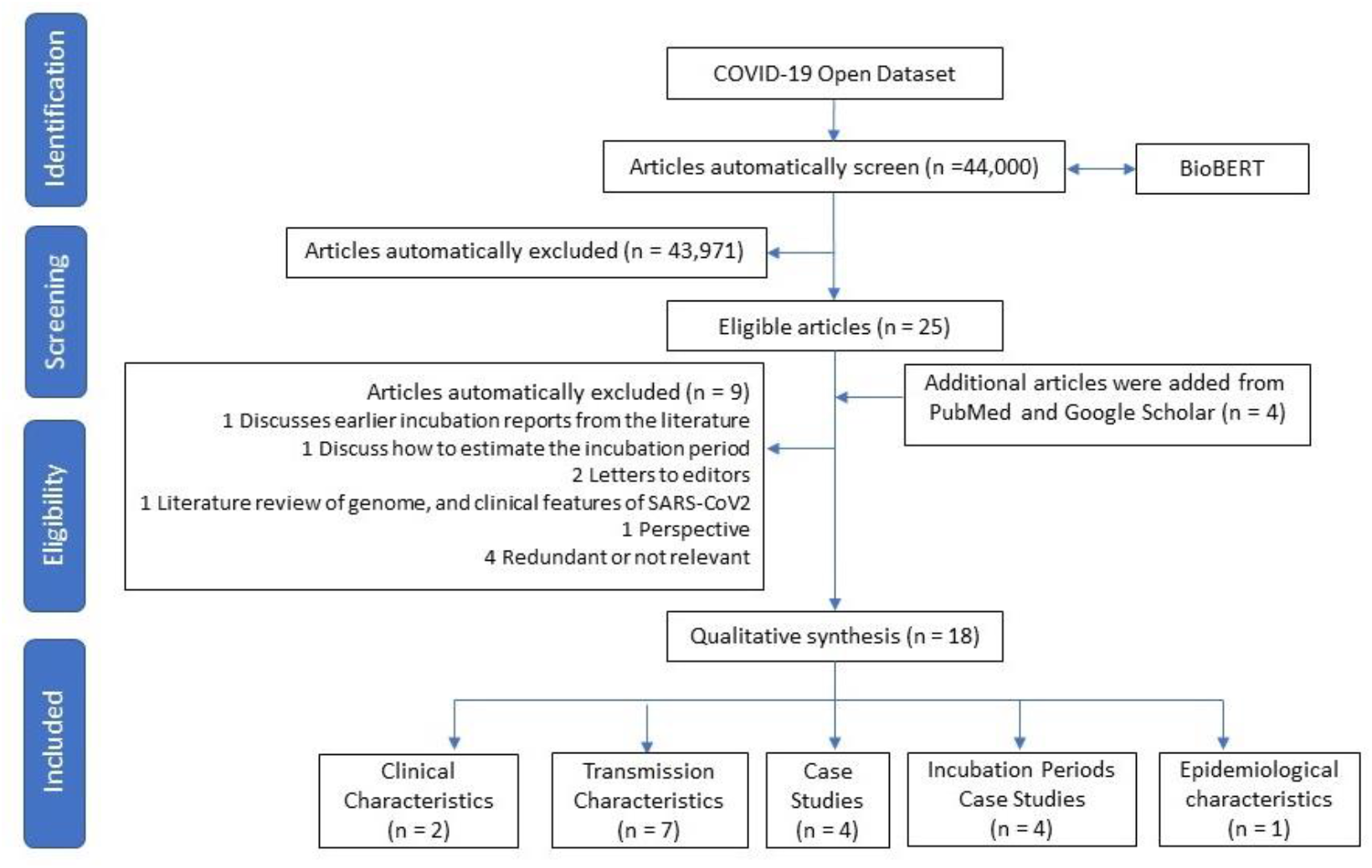
Flowchart of the study inclusions and exclusions of articles.

### 3.2. Synthesis of Results

#### 3.2.1. Estimating the incubation period

Backer J.et al [1) employed travel histories and symptom onsets for 88 confirmed cases discovered beyond the boundaries of Wuhan, China, in the early stage of the coronavirus outbreak. These authors estimated that the incubation period ranged from 2.1 to 11.1 days (2.5^th^ to 97.5^th^ percentile) and that the mean incubation period was 6.4 days (95% CI: 5.6-7.7). This research offers empirical evidence and falls into the previously mentioned incubation periods estimated by both the ECDC and the WHO [11]. The researchers employed three parametric forms related to the distribution of the incubation period: lognormal distribution, gamma distribution, and the Weibull distribution. They employed uniform prior probability distribution for the exposure interval related to the point of infection for all 88 individuals. The researchers used posterior distribution samples employing the RStan package within R software version 3.6.0 (R Foundation, Vienna, Austria) [12].

Natalie ML. Et al [2] examined COVID-19’s epidemiological characteristics and incubation period. The researchers harvested information relating to confirmed diagnoses of COVID-19 infection beyond the disease epicentre in Wuhan, China, using official reporting from state institutes and reporting on mortalities both within and outwith Wuhan. The data used by the authors was either directly harvested from government sources or from news websites reporting government statements. The data collection process was real-time, so it was added to as further details emerged. The final data selection represented a selection of reported cases up to January 31, 2020. The outcomes of this research concluded that the incubation period falls into a range of 2-14 days (95% CI), with the mean being approximately five days as found by employing best-fit lognormal distribution. Mean time between the onset of symptoms and admission to hospital (either for treatment or isolation) was estimated to be between three and four days with no truncation and between five and nine days with right truncation. On the basis of the 95^th^ percentile estimate for the incubation period, the researchers recommended that exposed individuals should be quarantined for a minimum of 14 days. When making estimates of the risk of fatality in COVID-19 cases, the median time delay between illness onset and death of 13 days (17 days with right truncation) should be taken into account.

Jiang X et al [13] undertook research making a comparison between incubation periods for MERS, SARS, and SARS-CoV-2. The researchers reported that SARS-CoV-2 has an extended incubation period, which has led to modifications in official policy for control and screening. To prevent the virus spreading, any individual who may have been exposed should go into isolation for 14 days, this being the outer limit of predictions for incubation times. Nevertheless, by analyzing a large dataset for this research, researchers report that no identifiable difference exists between incubation times for SARS-CoV-2, severe acute respiratory syndrome coronavirus (SARS-CoV), and the Middle East respiratory syndrome coronavirus (MERS-CoV), which highlights the requirement for more extensive and better-annotated datasets. This research covered 49 patients with SARS-CoV-2 who had definite dates for the first exposure, end of exposure and beginning of symptoms, 153 patients with SARS-CoV, and 70 MERS-CoV patients; this data was amalgamated from seven separate papers. The results indicated that MERS incubates on average 5.8 days (95% CI: 5-6.5), SARS-CoV 4.7 days (95% CI: 4.3-5.1), and SARS-CoV2 4.9 days (95% CI: 4.4-5.5). This demonstrates that the longest incubation period is MERS-CoV, with SARS-CoV2 second longest.

Lauer Stephen et al [14] researched the COVID-19 incubation period by looking at diagnosed cases that have been publicly reported. The aim of the study was to ascertain COVID-19’s incubation period and to detail its implications for public health. The researchers examined diagnosed cases of COVID-19 occurring between January 4, 2020, and February 24, 2020. The research covered 181 subjects diagnosed with SARS-CoV-2 infection outwith Hubei province, China by examining press releases and news reports from 50 different provinces, regions, and nations. The researchers harvested information regarding patient demographics, dates/times of possible exposure, the onset of symptoms, the onset of fever, and admission to the hospital. The researchers estimate that, conservatively, 101/10,000 cases (99^th^ percentile, 482) will experience symptom onset more than 14 days after being quarantined or actively monitored. Nevertheless, the researchers noted that severe cases could be overrepresented in public reporting; it is possible that severe and mild COVID 19 infections have different incubation periods. This research adds to the evidence that COVID-19 is similar to SARS in having a median incubation period of around five days. This recommendation comes from the research looking at proposed quarantine/active monitoring times for subjects with potential exposure to the virus.

#### 3.2.2. Transmission Characteristics

Li Q. et al [15] researched the early data regarding transmission dynamics for the virus in Wuhan, estimating the mean incubation period at 5.2 days (95% CI: 4.1-7.0), with the distribution’s 95^th^ percentile being 12.5 days. The researchers fitted a log-normal distribution to data regarding the history of exposure and date of onset using only those cases where detailed information was available to estimate the length of incubation. Their preliminary estimate of the distribution of the incubation period offers strong support for the case that exposed subjects should be quarantined or put under medical observation for 14 days. Nevertheless, this study’s accuracy may be questioned as the estimate was made using data from just 10 patients.

Meili L. Et al [16] researched the transmission characteristics of China’s COVID-19 outbreak. They took individual patient histories from COVID-19 subjects in China (not from Hubei Province) for estimating the distribution of the time for generation, incubation, and the time span between onset of symptoms and isolation/diagnosis. On average, patients were isolated 3.7 days after the onset of symptoms, and diagnosed after 6.6 days. The average patient was found to be infectious 3.9 days prior to displaying any notable symptoms. This militates against effective quarantining or contact tracing. With contact tracing, isolation, and quarantine, the baseline reproduction number is estimated at 1.54, with the majority of infection attributable to super spreaders. The authors of this research, on this basis, suggested that 14 day quarantine periods would fail for 6.7% of subjects; they proposed 22-day quarantine becoming standard. This research gave an estimation of the mean incubation period as 7.2 days. Lauren C.T.et al [17] researched the estimations for transmission intervals for COVID-19. The researchers used transmission clusters for estimations of serial interval distribution and incubation period. They used information on outbreaks in Tianjin, China (between January 21 and February 27), and Singapore (between January 19 and February 26). Interval censoring was employed (R package icenReg [18]) for making parametric estimations of the distribution of the incubation period. Mean incubation periods for Tianjin were estimated at nine days (7.92, 10.2) and for Singapore 7.1 days (6113, 8.25). It was additionally recorded by the researchers that in both datasets cases that occurred earlier showed shorter incubation periods.

China Chengfeng Q. et al [19] undertook research into the virus’s transmission and clinical characteristics. This research comprised a contact investigation involving 104 patients in special hospitals in Hunan province from January 22, 2020, and February 12, 2020. Collection and analysis were made of information regarding patient demographics, clinical, laboratory, and radiological findings, medication administered, and patient outcomes. Confirmation of patient illness was made with the PCR test. The patients had a mean age of 43 (ranging from 8 to 84), with 52.88% female. The researchers found a median incubation period of six (range 1-32) days; eight patients incubated between 14 and 17 days, and eight patients incubated between 18 and 30 days.

Yan Bei et al [20] undertook research into cases where it was assumed that the virus had been passed by an asymptomatic carrier. This research looked at a family from Anyang, China, of which five members were suffering COVID-19 pneumonia who had, prior to developing symptoms, been in contact with an asymptomatic member of the family who had traveled to see them from Wuhan, the origin of the pandemic. The timeline they uncovered implies that the coronavirus could have been passed on by this asymptomatic carrier. The first patient to develop symptoms incubated for 19 days, a long period but one which falls within the reported range (0 to 24 days). This patient initially produced a negative return for the RT-PCR test; RT-PCR is a common test for diagnostic virology and does not often return false positives, so her second result from this test was probably not a false positive, and so it was assumed that she was infected with the coronavirus that is responsible for COVID-19.

Wei Xia et al [21] undertook research into how the coronavirus was transmitted in incubation periods in 2019. The researchers harvested data on the demographics, possible exposure, and time to symptom onset for confirmed cases published by local Chinese authorities. They assessed the possibilities of transmission in the course of the incubation period for 50 clusters of infection; these included 124 cases outwith Wuhan/Hubei province. Every secondary case examined and been in contact with a first-generation case prior to experiencing symptoms. This research found that the mean incubation period for COVID-19 was 4.9 days (95% CI, 4.4-5.4), with a range of 0.8 to 11.1 days (2.5^th^ to 97.5^th^ percentile). The infectious curve demonstrated that 73% of secondary cases became infected prior to symptoms appearing for first-generation cases; this was especially the case in the final three days of incubation. These findings demonstrated that COVID-19 is transmitted between those who are in close contact in the course of incubation, which could indicate a weakness in the quarantine system. Robust workable countermeasures are required for the prevention or mitigation of the virus being transmitted asymptomatically amongst high-risk populations in the course of the incubation period.

Juanjuan Z. [22] undertook research into the evolution of COVID-19’s transmission dynamics and epidemiology outwith Hubei province. It was hoped that being able to understand the evolution of the transmission dynamics and epidemiology outwith the center of the epidemic would provide useful information that could be used as guidance for intervention policies. The researchers harvested data on individuals whose diagnosis was confirmed by laboratory testing in mainland China (apart from Hubei) that was reported by official public sources between January 19 and February 17, 2020. The date of the fourth time the case definition was revised (January 27) was employed as a means of dividing the epidemic into two phases (December 24-January 27 and January 28-February 17) as dates for symptom onset. Trends were estimated in terms of central time-event periods and subject demographics. This research encompassed 8579 cases, covering 30 provinces. The median age of the subjects was 44 years (range 33 to 56); as the epidemic continued, more cases emerged in the younger age groups and for the elderly (those aged over 64). Mean time between symptom onset and hospitalization fell from 4.4 days (% 95% CI 0.0-14.0) between December 24 and January 27, down to 2.6 days (0.0-9.0) between January 28 and February 17. For the whole period, the mean incubation period was calculated at 5.2 days (1.8-12.4) with the mean serial interval being 5.1 days (1.3-11.6).

#### 3.2.3. Clinical Characteristics

Shaoqing et al [23] undertook research investigating the clinical characteristics and outcomes for patients submitting to surgery in the COVID-19 incubation period. The researchers undertook the analysis of clinical data for 34 subjects submitting to elective surgery during the COVID-19 incubation period at four Chinese hospitals (Renmin, Tongji, Zhongnan, and Central) in Wuhan between January 1 and February 5, 2020. The patients had a median age of 55, with 20 of them (58.8%) being female. Every patient exhibited COVID-19 pneumonia symptoms within a short time of surgery, with abnormalities showing on chest CT scans. Symptoms exhibited by these patients encompassed fever (31 (91.2%)), fatigue (25 (73.5%)) and a dry cough (18 (52.9%)). 15 of the patients (44.1%) had to be admitted to the intensive care unit (ICU) as the disease progressed, and seven of these died (20.5%). Every patient examined in this research had been directly exposed to the environment in Wuhan before being admitted to the hospital; no patient had exhibited any symptoms of COVID-19 prior to surgery. It was notable how swiftly the COVID-19 symptoms appeared once surgery had been completed, with the infection being confirmed by the laboratory within a short period. The time gap between hospital admission and surgery (median time 2.5 days) is less than the median incubation period of 5.2 days found in patients with laboratory-confirmed infections in Wuhan [15]; it is also less than the general incubation time in hospitals in China (median time 4.0 days, from research into infected patients from 552 Chinese hospitals [19]). Taken together, this evidence is confirmation of the hypothesis that patients within this research were incubating COVID-19 prior to submitting to surgery.

Guan et al [24] undertook research into coronavirus’ clinical characteristics within China. They reviewed data for 1099 patients who had a COVID-19 diagnosis confirmed by the laboratory; data came from 552 hospitals across 32 provinces and municipalities of mainland China up to January 29, 2020. Only data from 291 patients were used to calculate incubation periods, these being patients who had a clear idea of the date they had been exposed. The patients had a median age of 47 years, with 58.1% male and 41.9% female. 1.18% of the subjects had been in direct wildlife contact, 31.30% had visited Wuhan, and 71.80% had been in contact with people from Wuhan. There was found to be a median incubation period of four days (interquartile range between two and seven days).

#### 3.2.4. Case studies

Jasper F.C. et al [25] looked at a cluster of pneumonia within a family associated with COVID-19. The researchers examined clinical, laboratory, epidemiological, microbiological, and radiological outcomes for five patients (aged between 36 and 66 years) from the same family who all manifested idiopathic pneumonia upon their return to Shenzen, Guangdong province, having visited Wuhan from December 29, 2019, to January 4, 2020. All patients exhibited at least one and sometimes more of the symptoms of diarrhea, upper/lower respiratory tract problems, or fever, within 3 to 6 days after being exposed. The patients attended hospital between six and ten days from symptoms appearing. The findings of this research accord with other accounts of COVID-19 transmission in family/hospital environments, and share features with reports of travellers with the infection in other areas. Jin-Wei et al [26] undertook research into 102 cases of COVID-19 (51% male, 49% female) in Xiangyang, China, all of whom tested positive via RT-PCR. The cohort ranged in age from 1.5 years to 90 years, with a mean age of 50.38. The majority of subjects were aged between 50 and 70 years. Of the 71 subjects who had confirmed contact histories, 7 lived in Wuhan, 37 had traveled there, 4 had contact with patients who had been diagnosed with the virus, and 23 were members of families where infection clusters appeared. Analyzing 44 subjects where it was obvious where the contact occurred, incubation periods were between 1 and 20 days, with the mean being 8.09 days (4.99). Severe illness/death rates were lower than average, with certain patients incubating the virus for longer periods than would be predicted. The researchers made a recommendation that quarantine periods should be extended to 3 weeks where appropriate.

Yuanyuan H. l [27] undertook research into COVID-19 patients who were long-term users of glucocorticoids. Across the world, clusters of patients having COVID-19 have been reported, with research demonstrating that person-to-person transmission is the chief route of infection. The research states that average incubation periods are between 2 and 14 days, with between 3 and 7 days being the most common. The research notes that incubation periods may be extended in some patients. The research reports on a family cluster of COVID-19; a 47-year-old female member of the family on long-term glucocorticoid therapy had no symptoms in a 14 day quarantine period but tested positive for COVID-19 antibodies 40 days after leaving Wuhan. These findings imply that long-term glucocorticoid use could be responsible for long incubation periods, atypical infection, and additional transmissions of the virus.

Rachael P.F et al [28] undertook an investigation into Singapore’s surveillance/response measures. This research examined three COVID-19 clusters associated with a church in Singapore, a business conference, and a visiting Chinese tour group in February 2020. The research employed inpatient medical records and interviews to harvest clinical and epidemiological data regarding subjects with a confirmed diagnosis of COVID-19; field investigations were undertaken for assessment of the ways these subjects had interacted with others and the potential ways in which they had acquired the virus. With overseas cases, open-source reports were used. At the time of the research (February 15, 2020) there were 36 infections with epidemiological links to the three Singaporean clusters mentioned above. 425 subjects who had had close contact with people from these clusters were put in quarantine. Those affected had generally had prolonged direct close contact with others, although it was not possible to exclude indirect transmission, e.g. through shared food or fomites. This research found a median incubation period for COVID-19 of four days (IQR 3-6). Serial intervals for transmission pairs were between three and eight days. The researchers came to the conclusion that the virus can be transmitted within communities and that local clusters of infection will appear in countries that welcomed high numbers of Chinese visitors prior to Wuhan being locked down and travel restrictions being imposed. The research further concluded that contact tracing and increased surveillance is necessary to prevent the virus from spreading widely across communities.

#### 3.2.5. Epidemiological characteristics

China Pei W. et al [29] undertook an investigation into COVID-19’s epidemiological characteristics as found in the Chinese province of Henan, centered on publicly available data related to 1212 patients. The report mentions that these patients are 55% male and 45% female, with 81% being aged from 21 to 60 years. Statistical analysis undertaken with 483 patients on this cohort revealed that the estimated average median period was 7.4 days, the mode 4 days, and the median 7 days. 92% of patients did not have an incubation period in excess of 14 days.

### 3.3. Results of individual sources of evidence

Table 1 summarises all of the articles under review/analysis, including the methodology employed for the estimation of incubation periods.

**Table 1:** An overview of articles investigating and reporting on the incubation period of COVID-19.

| Ref. | Incubation Period | Method for estimating the incubation period. | Number of cases | Location | Note |
| --- | --- | --- | --- | --- | --- |
| **Incubation Period Studies** |  |  |  |  |  |
| [1] | 6.4 days (95% CI: 5.6–7.7), ranging from 2.1 to 11.1 days (2.5th to 97.5th percentile) | Probability density function (PDF). Used a doubly interval-censored likelihood function to estimate the parameter values. Used RStan package in R | 88 | Outside Wuhan, China | - The study provides empirical evidence and the estimation is within the range for the incubation period of 0 to 14 days assumed by the WHO and of 2 to 12 days assumed by the ECDC [11]. |
| [2] | 5 days (2–14 days with 95% confidence) | Best-fit lognormal distribution | Real-time data | outside of the Wuhan, China | - Recommend the length of quarantine to be at least 14 days.<br>- The median time delay of 13 days from illness onset to death should be considered when estimating the fatality risk. |
| [13] | - SARS-CoV2 4.9 (95% CI:4.4-.5.5)<br>- SARS-CoV 4.7(95% CI: 4.3-5.1)<br>- MERS-CoV 5.8 (95% CI: 5-6.5) | Fitted Weibull, lognormal, and gamma distributions | - SARS-CoV2 49<br>- SARS-CoV 153<br>- MERS-CoV 70 | China | - long incubation time was reported to be associated with SARS-CoV-2 infection, leading to adjustments in screening and control policies. |
| [14] | Median 5.1 days (95% CI, 4.5 to 5.8 days) | Estimated the incubation time using a previously described parametric accelerated failure time model. Doubly interval-censored data. Data reduction technique | 181 | Regions, and countries outside Wuhan, Hubei province, China | - 101 out of every 10,000 cases (99th percentile, 482) develop symptoms after 14 days of active monitoring or quarantine.<br>- This work provides additional evidence for a median incubation period of approximately 5 days, like SARS. |
| **Transmission Characteristics** |  |  |  |  |  |
| [15] | 5.2 days (95% confidence interval (CI): 4.1-7.0), with the 95th percentile of the distribution at 12.5 days | Estimated by fitting a log-normal distribution to data on exposure histories and onset dates in a subset of cases with detailed information available. | 10 | Wuhan, China | - The estimation was based on information from only 10 cases and is somewhat imprecise.<br>- The incubation period reported is Within the WHO limit. |
| [16] | Mean of 7.2 days | Gamma distribution and a log-normal distribution | - | Chinese provinces excluding Hubei | - The authors stated that the recommended 14-day quarantine period may lead to a 6.7% failure for quarantine and they suggested a 22-day quarantine period. |
| [17] | Mean incubation periods as<br>- 7.1 (6.13, 8.25) days for Singapore and<br>- 9 (7.92, 10.2) days for Tianjin | Used interval censoring R package icenReg [18] to make parametric estimates of the incubation period distribution | - 93 Singapore<br>- 135 Tianjin | Singapore, Tianjin (China) | - Both datasets had shorter incubation periods for earlier-occurring cases. |
| [19] | Median incubation period was<br>- 6 (rang, 1-32) days, of 8 patients ranged from 18 to 32 days. | Recorded Observation | 104 | Hunan, outside-Wuhan | - The incubation period of 8 patients exceeded 14 days. |
| [20] | 1-19 days | Observation | 5 | Anyang, China | - The incubation period for patient 1 was 19 days, which is long but within the reported range of 0 to 24 days |
| [21] | be 4.9 days (95% confidence interval [CI], 4.4 to 5.4) days, ranging from 0.8 to 11.1 days (2.5th to 97.5th percentile) | Used a Weibull distribution-based survival analysis model with the extension of the Kaplan-Meier estimator to fit the curve of the incubation period for COVID-19 cases. | 124 | Outside Wuhan city and Hubei province in China | - 73.0% of the secondary cases, their date of getting infected was before symptom onset of the first-generation cases, particularly in the last 3 days of the incubation period.<br>- Strong and effective countermeasures should be implemented to prevent or mitigate asymptomatic transmission during the incubation period in populations at high risk. |
| [22] | 5.2 days (1.8–12.4) and the mean serial interval at 5.1 days (1.3–11.6). | Lognormal distribution | 8579 cases from 30 provinces | outside Hubei in mainland China | - The mean time from symptom onset to hospital admission decreased from 4.4 days (95% CI 0.0–14.0) from Dec 24 to Jan 27, to 2.6 days (0.0–9.0) for the period of Jan 28 to Feb 17. |
| **Clinical Characteristics** |  |  |  |  |  |
| [23] | Median incubation period of 2.5 | Lognormal distribution | 34 | Wuhan | - The length of time from hospital admission to surgery (median time, 2.5 days is shorter than the median incubation time of 5.2 days |
| [24] | Median incubation period of 4 | Median, Observation (2-7 days) | 291 | Mainland China | - The median age was 47 years, and 41.90% were females. Only 1.18% of patients had direct contact with wildlife, whereas 31.30% had been to Wuhan and 71.80% had contacted people from Wuhan. |
| **Case Study** |  |  |  |  |  |
| [25] | - Patient 3 – (3-6 days after exposure)<br>- Patients 1-4 symptomatic (6-10 days after symptom onset)<br>- 5 not affected | Observation | 5 | Wuhan | - 5 patients (aged 36–66 years)<br>- The authors' findings are consistent with person-to-person transmission of the novel coronavirus in hospital and family settings, and the reports of infected travelers in other geographical regions. |
| [26] | Mean of 8.09 (4.99) days | Mean, Observation | 44 | Xiangyang, China | - Most cases fell into the age group of 50-70 years old.<br>- The rate of severe illness and death was low, whereas some patients had a longer incubation period. |
| [27] | 2–14 days, and mostly 3–7 days. | Observation | 3 | Hunan, China | - A 47-year-old woman with long-term use of glucocorticoids did not develop any symptoms within the 14-day quarantine period.<br>- The results suggest that the long-term use of glucocorticoids might cause atypical infections, a long incubation period, and extra transmission of COVID-19.<br>- Case 1 is a Wuhan-settled 47-year-old female. She has a more than 16-year history of systemic lupus erythematosus (SLE)<br>- Case 2 is an 81-year-old male who lives in Yiyang. He has a history of prostate cancer and coronary heart disease.<br>- Case 3 is a 44-year-old female from Yiyang. She also had close contact with her elder sister (Case 1). |
| [28] | The Median incubation period is 4 days. | Observations | 36 | Tour group from China, in Singapore | - Interpretation SARS-CoV-2 is transmissible in community settings, and local clusters of COVID-19 are expected in countries with high travel volume from China.<br>- Enhanced surveillance and contact tracing are essential to minimize the risk of widespread transmission in the community. |
| **Epidemiological Characteristics** |  |  |  |  |  |
| [29] | Estimated average, mode and median incubation periods are 7.4, 4 and 7 days, respectively | log-normal distribution | 483 | Henan, China | - COVID-19 patients show gender (55% vs 45%) and age (81% aged between 21 and 60) preferences, possible causes were explored.<br>- The incubation periods of 92% of the patients did not exceed 14 days. |

## 4. Discussion

### 4.1. Summary of evidence

In this Scoping Review, we identified 18 primary studies estimated the length of incubation periods among COVID-19 patients published between December 1, 2019, and April 25, 2020. On the basis of this research, it appears that the only means of estimating the incubation period for COVID-19 at present is logarithm normal distribution. Novel ways of effecting improvements to the estimation’s accuracy in terms of estimating incubation periods should be investigated. This is an important issue because the date of infection and/or onset of symptoms are difficult to precisely identify. Nicholas G.R.et al [31] have made a comparison of two means of making such estimates. One method uses doubly interval-censored data, and the other employs data reduction techniques making the calculation more tractable. The researchers used both methods on historical data relating to the incubation periods for respiratory syncytial virus and influenza A. The outcomes demonstrate that these methods reduce the demands for computational power to analyze the reduced data and make good estimates of median incubation times in many different experimental conditions. Nevertheless, the researchers do recommend that doubly interval-censored analysis should be used to estimate the distribution tails.

The systematic review undertaken by this research has demonstrated that the currently published estimations have offered empirical evidence that the virus’ incubation period is approximately a median of 5.01 and a mean of 7.8 days, which falls into the 2 to 12 days assumption of the ECDC and the 0 to 14 days of the WHO [11].

Researchers like [2] propose a recommended quarantine length of a minimum of 14 days, suggesting that fatality risks should be estimated using a median time of 13 days between first symptoms and mortality. [14] offered evidence that it is possible for symptoms to appear once patients have left a 14 day period of quarantine or active monitoring; the authors suggest that the median incubation time is around five days, similar to SARS. In [16], the authors stated that 14-day quarantines may be insufficient to protect the public in 6.7% of cases; they, therefore, proposed that quarantine should be a minimum of 22 days. In [19] it was found that eight patients developed symptoms after more than 14 days from infection. In [20] the authors referred to a patient who incubated the virus for 19 days, a substantial period but one which falls into the reported range (0-24 days).

It has yet to be conclusively demonstrated whether the age group as any impact on the time a patient incubates the virus. Out of 291 patients [24] with an average age of 47, the incubation period was 4.0 days, for five patients [25] with an average age of 49.5 years it was 4.5 days, for 44 patients [26] with an average age of 60 years it was 4.99 days, and for two patients [27] with an average age of 47 years, it was 4.5 days.

The authors of [27] suggested that long-term glucocorticoid use could cause a delay in the onset of symptoms so that they would not appear until after a 14 day quarantine period had been completed. Long-term glucocorticoid use could be responsible for the atypical infection, longer incubation periods, and additional COVID-19 transmission.

[13] states that COVID-19 has a mean incubation period lower than MERS coronavirus but higher than SARS coronavirus. [21] offers a final insight that robust and workable countermeasures must be put in place for the prevention and/or mitigation of asymptomatic transmission in the course of incubation amongst high-risk populations.

### 4.2. Limitations

The Scoping Reviews presented in this paper has some limitations. The reviews included not peer-reviewed articles. No meta-analysis was performed. Furthermore, this Scoping Review was an enormous undertaking and our results are only up to date as of April 25th, 2020. This research has looked at a single element of the virus behavior, viz. its incubation period. While health authorities must know how they can accurately estimate the virus’ incubation period in order to direct the most effective public health interventions, select the best course of action and make estimations of the size of the epidemic, there is still no definitive answer as to what the incubation period is.

### 4.3. Conclusions

Throughout human history, epidemics have been a disrupter of human civilizations and caused staggering amounts of mortality and illness for both humans and animals [30]. A novel form of coronavirus (Sars-CoV-2) appeared in Wuhan, China, in December 2019; the virus was unexpected and swiftly spread worldwide. Many leading research laboratories and the WHO are striving to create a vaccine to protect against the disease. Nevertheless, as this is a new form of virus, there is much still to understand it. It is essential that we gain an understanding of the way the disease behaves.

In conclusion, the majority of extant published articles estimates offer empirical evidence showing that the incubation period for the virus is a mean of 7.8 days, with a median of 5.01 days, which falls into the ranges proposed by the WHO (0 to 14 days) and the ECDC (2 to 12 days). Moreover, a number of authors proposed that quarantine time should be a minimum of 14 days and that for estimates of mortality risks a median time delay of 13 days between illness and mortality should be under consideration.

It is unclear as to whether any correlation exists between the age of patients and the length of time they incubate the virus. Finally, it is generally agreed that robust precautions must be put in place for the prevention and/or mitigation of asymptomatic transmission to high-risk patients caused by those incubating the virus.

## Data Availability

This is a literature review paper.

## Acknowledgment

The authors would like to acknowledge the encouragement and support provided by Ajman University and United Arab Emirates University for conducting this research work.

## Ethical approval

Not required.

